# Closing the Gap: A Binational Analysis of Diabetes Mortality and Disability across the USA-Mexico Border Region

**DOI:** 10.1101/2025.10.28.25337683

**Authors:** Kaustubh Wagh, Jessica L Harding, Carlos A. Aguilar Salinas, Kathryn E. Kanzler, Gerardo Chowell

## Abstract

**Aims:** This study characterizes the diabetes burden across USA–Mexico border states (1990–2021) by examining current patterns, temporal trends, binational comparisons, and demographic variations.

**Methods:** Using Global Burden of Disease 2021 estimates, we calculated age-standardized rates for mortality, prevalence, incidence, and DALYs. We applied joinpoint regression, age-period-cohort models, and decomposition analyses to characterize trends and identify drivers of burden change, comparing border versus non-border states within and between countries.

**Results:** Border states showed 9-14% higher burden than non-border states. Despite similar prevalence (9,010 vs 8,298 per 100,000) and incidence (435 vs 426 per 100,000), Mexican border states had nearly five times higher mortality (67.2 vs 12.0 per 100,000) and three times higher DALYs (2,355 vs 896 per 100,000) than USA counterparts. YLLs comprised 64.3% of Mexican burden versus 31.4% in USA. From 1990-2021, USA border states showed rising incidence (+3.04% annually) with declining mortality (- 1.28%), while Mexican states demonstrated stable-to-declining incidence (-0.15%) with plateauing mortality post-2009. Males experienced higher mortality in both countries.

**Conclusions:** Diabetes occurrence has converged across the border, yet outcomes remain profoundly unequal. This demands coordinated binational surveillance and policy action. Focus on prevention in the US and diabetes care management in Mexico to reduce health disparities.

## 1. Introduction

Diabetes mellitus is one of the most significant global health challenges of the 21st century, affecting over 589 million people worldwide and causing approximately 3.4 million deaths annually (Magliano et al., 2025). As a leading cause of blindness, kidney failure, heart attacks, stroke, and lower limb amputation (WHO, 2016), the burden of diabetes is unevenly distributed across populations. This is particularly evident in the Americas where diabetes prevalence increases over the past three decades, driven by population aging, urbanization, dietary transitions, and rising obesity (Gakidou et al., 2011; Zhou et al., 2016). Mexico (22.8%) and the United States (21.9%) have the one of the highest proportions of diabetes-related deaths globally among people aged 20-79, far exceeding all other regions and signaling a severe public-health crisis in North America (Magliano et al., 2025). In border states, shared risk factors and cross-border movement create unique epidemiological patterns characterized by interrupted healthcare access, rapid dietary transitions toward processed foods, and barriers to consistent disease monitoring across health systems (Barquera et al., 2018; Magliano et al., 2025).

The USA-Mexico border region spans approximately 2,000 miles across six Mexican states (Baja California, Sonora, Chihuahua, Coahuila, Nuevo León, and Tamaulipas) and four USA states (California, Arizona, New Mexico, and Texas) (Border Health Commission, 2024). Border populations face elevated diabetes risk, with prevalence rates two to three times higher than non-border populations (Fisher-Hoch et al., 2010; PAHO, 2010). This stems from the convergence of multiple socioeconomic and environmental factors unique to border regions like extreme obesity rates exceeding 57% in some border communities, concentrated poverty with median household incomes 20-30% lower than national averages, limited healthcare infrastructure, food insecurity, unique stressors related to immigration status, and cross-border mobility (Power & Byrd, 1998; Homedes, 2012; CDC, 2024). Researchers describe a “the tip of the iceberg” phenomenon, where diagnosed cases represent only a fraction of the true burden due to high undiagnosed rates and limited healthcare access (Bastida et al., 2008; PAHO, 2010). The predominantly Hispanic/Latino population experiences diabetes prevalence rates 50-70% higher than non-Hispanic whites, with earlier onset and more severe complications (Pérez-Escamilla, 2011; Fisher-Hoch et al., 2010). While the USA-Mexico border region is traditionally defined as 100km north and south of the international boundary, measuring burden at the state level is valuable because health policies operate through state administrative structures (Antonio-Villa et al., 2024), transnational factors extend throughout border states (Barquera et al., 2018), and state-level data provide necessary statistical power (Mardon, 2017), and GBD data are available at the state level enabling standardized binational comparisons.

Epidemiological research documents diabetes prevalence rates of 12-18% among Hispanic adults in border communities, substantially higher than the 8-10% in non-border regions (Fisher-Hoch et al., 2010; Reininger et al., 2015; Gregg et al., 2018; Barquera et al., 2013). However, existing studies focus on individual communities or single states using varied methodologies, limiting regional generalizability. Cross-border health research faces significant challenges related to data harmonization and differing surveillance systems, which previous studies have address through national survey data often lacking subnational granularity and comparability (Díaz-Apodaca et al., 2010; Diaz-Kenney et al., 2010; Homedes & Ugalde, 2003; McEwen et al., 2009; Warner, 1991). While the Global Burden of Disease (GBD) framework offers standardized methodology for cross-national disease burden assessment, its application to subnational transnational analysis remains underutilized (Murray & Lopez, 2013). This study addresses key limitations like absence of comprehensive, standardized analysis across all ten border states; lack of binational comparative analysis using consistent methodology; unexplored evolution from mortality-focused to disability-focused disease burden; and inconsistent demographic pattern analysis. This is the first harmonized, long-term analysis of diabetes burden (1990–2021) across all ten USA–Mexico border states using GBD estimates. Our objectives are to: (1) characterize current diabetes burden across border states using 2019-2021 data, including mortality, prevalence, incidence, years of life lost (YLL), years lived with disability (YLD), and disability-adjusted life years (DALYs); (2) analyze three-decade trends; (3) compare burden patterns between the two countries and assess convergence or divergence; (4) examine demographic variations by age and sex. We hypothesize that: (1) significant border vs non-border states diabetes burden will exist within each country; (2) diabetes burden will show increasing trends over time in both countries, with disability-related metrics rising more rapidly than mortality; (3) Mexican border states will demonstrate higher age-standardized diabetes burden; and (4) demographic patterns will reveal higher burden among older adults, with potential sex differences in disease outcomes.

## 2. Materials and Methods

### 2.1 Study Design and Setting

This study employed a cross-sectional and longitudinal comparative analysis of diabetes burden across USA-Mexico border states from 1990-2021. The analysis focused on six Mexican border states (Baja California, Sonora, Chihuahua, Coahuila, Nuevo León, and Tamaulipas) and four USA border states (California, Arizona, New Mexico, and Texas) with combined populations exceeding 100 million people. This state-level approach extends beyond the traditional 100km border corridor (which encompasses approximately 17 million residents) to capture the full scope of border-state populations and the epidemiological patterns that align with state boundaries rather than geographic proximity to the border (Border Health, 2024; US Census Bureau, 2024).

### 2.2 Data Sources and Preparation

#### 2.2.1 IHME Global Burden of Disease (GBD) 2021 Dataset

We assembled a state-level analytical dataset for diabetes burden across border states within the USA and Mexico using the Institute for Health Metrics and Evaluation’s Global Burden of Disease 2021 (GBD 2021) estimates for 1990–2021 (Global Burden of Disease Collaborative Network, 2024). From GBD 2021 we extracted count and rate for deaths, incidence, prevalence, disability-adjusted life years (DALYs), years of life lost (YLLs), and years lived with disability (YLDs), sex (male, female), 5-year age groups along with all age data. For comparability, analyses used age-standardized rates per 100,000 population with 95% confidence intervals, which allow valid cross-border and temporal comparisons by accounting for differences in population age structure across states and over time (Global Burden of Disease Collaborative Network, 2024).

### 2.3 Analysis

We followed analysis structure: (1) an overall summary of burden for the USA border states (4: California, Arizona, New Mexico, Texas) versus non-border states (rest of the USA) and Mexican border states (6: Baja California, Sonora, Chihuahua, Coahuila, Nuevo León, Tamaulipas) vs non-border states (rest of the Mexico), and (2) Comparison of border states of Mexico and USA including state-level analyses. We derived age- and sex-specific data for non-border estimates by subtracting border data from national data. We calculated GBD age-standardized rates with 95% confidence intervals for both annual (1990-2021) and multi-year period (2019-2021) stratified by sex and region type (border, non-border). Population estimates were obtained from counts and rates. For age standardization, we pooled age-specific data within each region type then applied GBD standard population weights.

#### 2.3.1 Current burden (2019-2021)

We quantified the current diabetes burden across border states within USA and Mexico using GBD 2021 estimates for “Diabetes mellitus” by sex and standard 5-year age groups. To stabilize estimates and mitigate single-year volatility (including pandemic-related concerns), we summarized 2019–2021 values as a three-year mean. Primary comparisons used age-standardized rates (per 100,000; GBD world standard population) to remove differences in age structure across states and sexes, ensuring like-for-like comparisons; age-specific rates were also examined to describe life-course patterns. For each metric (prevalence, incidence, deaths, DALYs, YLLs, YLDs), we calculated border-to-non-border ratios and Mexico-to-USA border ratios. Confidence intervals for ratios were computed using the delta method. Beyond direct rate comparisons, we calculated burden composition metrics by expressing YLLs and YLDs as percentages of total DALYs, and computed mortality-to-prevalence ratios (×10^−^³) to assess disease severity. Sex-specific analyses included calculating male-to-female ratios for each burden measure and region. Table 1 reports results stratified by country (Mexico, USA), region type (border, non-border), sex (male, female, both).

**Table 1.**
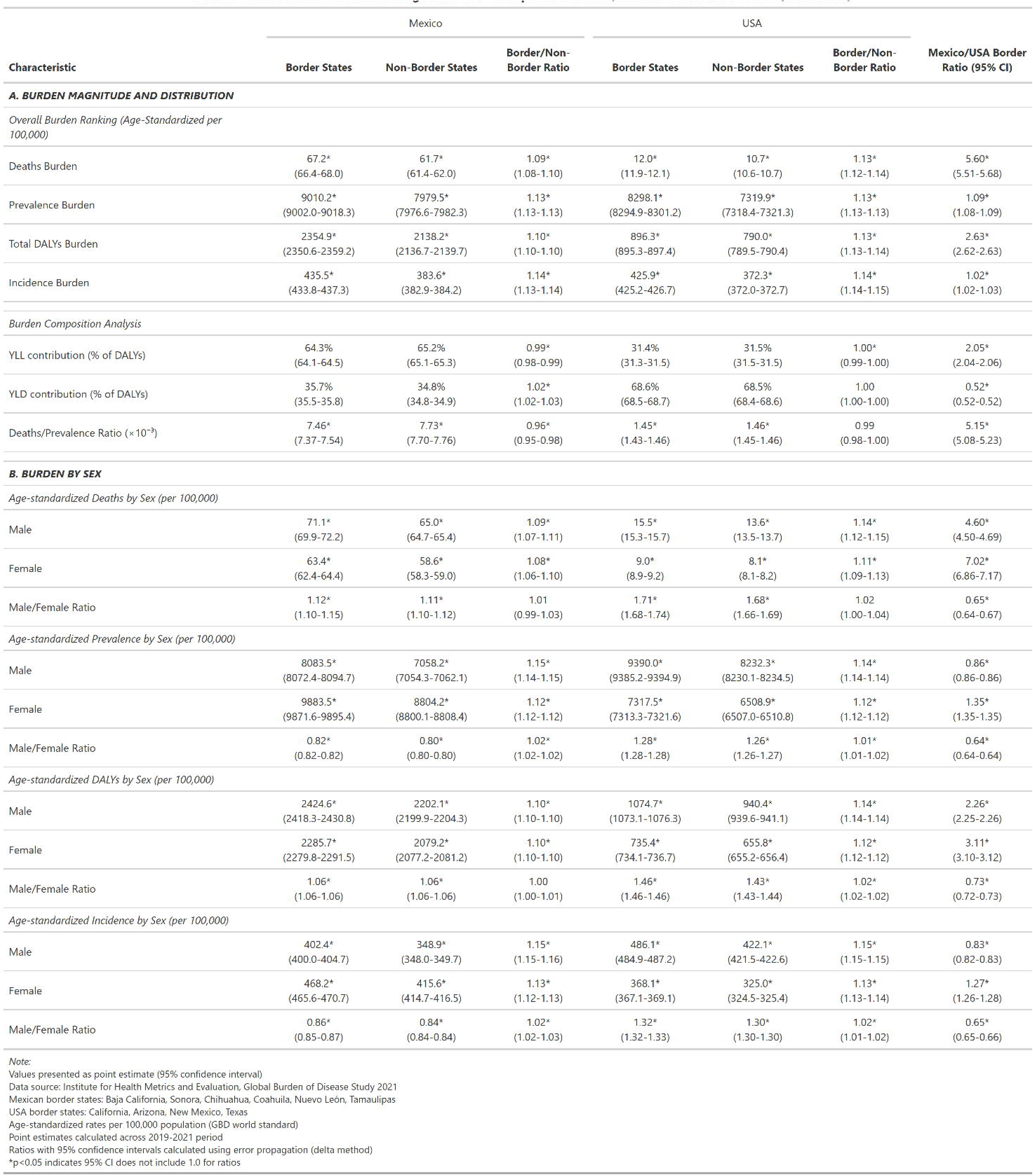
Current Diabetes Burden: Magnitude and Sex-Specific Pattens, Mexico-USA Border States (2019-2021)

#### 2.3.2 Trend Analysis (1990-2021)

We assessed temporal trends in age-standardized diabetes burden (deaths, prevalence, DALYs, incidence) from 1990 to 2021. We calculated average annual percent change (AAPC) for 1990-2021 using log-linear regression (ln(rate) = β + β ×year), with AAPC = 100×(exp(β) - 1) and 95% confidence intervals from regression coefficient limits. To test whether trends differed between groups, we added interaction terms: ln(rate) = β + β ×year + β ×group + β ×(year×group), where β tests if slopes differ. We compared border versus non-border trends within each country, Mexico border states versus USA border-states, and male versus female trends within each geography, yielding 52 total tests. All p-values were adjusted using Benjamini-Hochberg False Discovery Rate (FDR) correction, with significance defined as FDR-adjusted p < 0.05.

#### 2.3.3 Joinpoint Regression (1990-2021)

To characterize temporal trends in diabetes burden across border states within the USA and Mexico from 1990–2021, we fit joinpoint (segmented) log-linear regressions to annual age-standardized rates (per 100,000; GBD 2021, both sexes) for deaths, incidence, prevalence, and DALYs. Models were estimated on the natural-log of the rate so that segment slopes are interpretable as Annual Percent Change (APC = 100×[exp(β)−1])(CDC, 2025; Clegg et al., 2009; Geiss et al., 2014). We allowed up to two joinpoints and selected their number and locations using an information-criterion search that balances fit and parsimony. For each segment we report APCs with 95% confidence intervals; effects were considered statistically significant when the 95% CI excluded zero (denoted by an asterisk in the figure). We also computed an Average APC (AAPC) over the full interval as a duration-weighted geometric mean of segment APCs (Ahmed et al., 2025; NCI, 2025; Rojas-Martínez et al., 2024). Age standardization used the GBD world standard to ensure comparability across countries and over time. Figures show observed annual rates (points), fitted segments (solid lines), and estimated change-points (vertical dashed lines)

#### 2.3.4 Age-Period-Cohort Effect (1990-2021)

We quantified age, period, and cohort-specific effects in diabetes burden across border states within USA and Mexico using an age–period–cohort (APC) framework applied to IHME-GBD 2021 estimates for 1990–2021 (both sexes) (Columbia University, 2016; J. Liu et al., 2025). For DALYs, deaths, incidence, and prevalence, we aggregated state-level numerators and denominators within each country, derived birth cohort as calendar year minus age-group midpoint (<5 through 95+), and fit separate Poisson log-linear APC models with log population as an offset. To address the APC identifiability constraint, deviation (sum-to-zero) constraints were used, and exponentiated coefficients are reported as rate ratios relative to the grand mean for each effect (Rojas-Martínez et al., 2024). Uncertainty was quantified via 1,000 nonparametric bootstrap resamples of age–year strata within each country, with 95% CIs from the percentile distribution; country-specific age, period, and cohort rate ratios with uncertainty ribbons are shown in the figure.

#### 2.3.5 Decomposition Analysis (1990-2021)

We quantified drivers of change in the diabetes burden from 1990–2021 across border states within the USA and Mexico using IHME GBD 2021 estimates stratified by sex and harmonized 5-year age groups (<5 to ≥95 years) (C. Liu et al., 2025). For absolute numbers (deaths, prevalence, incidence, DALYs), we applied the N-component Das Gupta decomposition to partition total change into additive, order-invariant contributions from (i) population growth, (ii) population aging (age-structure shifts), and (iii) epidemiologic change (age-specific rate change) (Gupta, 1993; Yang et al., 2025). For crude rates (per 100,000), we used Kitagawa’s symmetric two-factor decomposition to attribute differences to (i) age-composition change and (ii) age-specific rate change (Kitagawa, 1955). To examine temporal heterogeneity, all decompositions were performed considering early period (1990–2005) and a recent period (2006–2021), using identical age groups to ensure comparability across places and sub-periods.

All statistical analyses were conducted using R version 4.3.0 with packages including tidyverse for data manipulation, mgcv for trend modeling, and ggplot2 for visualization (R Core Team, 2025).

This study utilized publicly available, de-identified aggregate health data and did not require institutional review board approval. All data use agreements and terms of service for the GBD database were followed.

## 3. Results

### 3.1 Current Burden (2019-2021)

Mexican border states had higher diabetes burden than non-border states across all measures (Table 1). Age-standardized death rates were 67.2 versus 61.7 per 100,000 (border/non-border ratio: 1.09, 95% CI: 1.08-1.10), prevalence was 9010.2 versus 7979.5 per 100,000 (ratio: 1.13), total DALYs were 2354.9 versus 2138.2 per 100,000 (ratio: 1.10), and incidence was 435.5 versus 383.6 per 100,000 (ratio: 1.14, 95% CI: 1.13-1.14). USA border states showed comparable patterns with border/non-border ratios of 1.13 for death rates, prevalence, and DALYs, and 1.14 for incidence. Sex-specific analyses revealed similar border effects for males and females in both countries.

Despite similar prevalence between Mexican and USA border states (9010.2 vs. 8298.1 per 100,000; ratio: 1.09), Mexican border states exhibited five-fold higher mortality (67.2 vs. 12.0 per 100,000; ratio: 5.60, 95% CI: 5.51-5.68) and nearly three-fold higher total DALYs (2354.9 vs. 896.3 per 100,000; ratio: 2.63, 95% CI: 2.62-2.63) (Table 1). Incidence rates were nearly identical (435.5 vs. 425.9 per 100,000; ratio: 1.02). Burden composition differed markedly between countries. YLL comprised 64.3% of DALYs in Mexican border states versus 31.4% in USA border states, while YLD accounted for 35.7% versus 68.6%, respectively. The deaths/prevalence ratio was 7.46 per 1,000 in Mexican border states compared to 1.45 per 1,000 in USA border states.

Within border states, males had higher death rates than females in both Mexico (71.1 vs. 63.4 per 100,000; ratio: 1.12, 95% CI: 1.10-1.15) and the USA (15.5 vs. 9.0 per 100,000; ratio: 1.71, 95% CI: 1.68-1.74) (Table 1). However, females had higher prevalence in Mexican border states (9883.5 vs. 8083.5 per 100,000 for males), while USA border states showed similar prevalence by sex (9317.5 vs. 9390.0 per 100,000). The Mexico/USA death rate ratio was 7.02 (95% CI: 6.86-7.17) for females compared to 4.60 (95% CI: 4.50-4.69) for males. Total DALYs followed the same pattern, with ratios of 3.11 (95% CI: 3.10-3.12) for females and 2.26 (95% CI: 2.25-2.26) for males.

In both USA and Mexico border states, diabetes burden increased substantially with age, with deaths concentrated in those aged 75 years and older (Supplementary Figure 1). Diabetes incidence peaked sharply in the 55-69 year age group in USA border states, whereas Mexican border states showed a broader distribution across ages 40-75, despite higher overall prevalence rates. Females demonstrated higher prevalence rates across most age groups in border states, whereas males experienced higher mortality rates, particularly among older adults.

### 3.2 Trend Analysis (1990-2021)

In Mexico, border states consistently outperformed non-border states across all metrics (all p<0.05, Table 2). Border states showed declining DALYs (AAPC = -0.62%, 95% CI: -0.77, -0.48) and incidence (AAPC = -0.15%, 95% CI: -0.22, -0.09), while non-border states had minimal DALY changes and rising incidence (AAPC = 0.18%, 95% CI: 0.12, 0.23) (Table 2). Conversely, USA border states demonstrated significantly higher increases in incidence and prevalence than non-border states (both p<0.05), though DALY and mortality trends did not differ significantly.

**Table 2.** Average Annual Percentage Change in Diabetes Burden (1990-2021)

| Sex | Mexico Border | Mexico Non-Border | Mexico B/NB Sig. | USA Border | USA Non-Border | USA B/NB Sig. | Mexico/USA Border Sig. |
| --- | --- | --- | --- | --- | --- | --- | --- |
| **DALYs** |  |  |  |  |  |  |  |
| Both | -0.62% (-0.77, -0.48) | -0.08% (-0.28, 0.12) | * | 1.16% (0.93, 1.39) | 1.04% (0.86, 1.22) |  | * |
| Male | -0.17% (-0.31, -0.04) | 0.32% (0.11, 0.52) | * | 1.43% (1.21, 1.66) | 1.27% (1.08, 1.45) |  | * |
| Female | -1.05% (-1.21, -0.90) | -0.44% (-0.65, -0.24) | * | 0.80% (0.56, 1.03) | 0.73% (0.55, 0.91) |  | * |
| Male/Female Sig | * | * |  | * | * |  |  |
| **Deaths** |  |  |  |  |  |  |  |
| Both | -0.97% (-1.17, -0.76) | -0.23% (-0.51, 0.05) | * | -1.28% (-1.83, -0.74) | -1.37% (-1.80, -0.93) |  |  |
| Male | -0.19% (-0.37, -0.01) | 0.49% (0.24, 0.74) | * | -0.69% (-1.21, -0.16) | -0.82% (-1.24, -0.39) |  |  |
| Female | -1.71% (-1.94, -1.48) | -0.90% (-1.21, -0.59) | * | -2.05% (-2.63, -1.47) | -2.06% (-2.52, -1.60) |  |  |
| Male/Female Sig | * | * |  | * | * |  |  |
| **Incidence** |  |  |  |  |  |  |  |
| Both | -0.15% (-0.22, -0.09) | 0.18% (0.12, 0.23) | * | 3.04% (2.93, 3.16) | 2.76% (2.64, 2.87) | * | * |
| Male | -0.20% (-0.27, -0.13) | 0.12% (0.05, 0.19) | * | 3.10% (2.99, 3.21) | 2.81% (2.70, 2.91) | * | * |
| Female | -0.11% (-0.18, -0.05) | 0.21% (0.16, 0.27) | * | 2.98% (2.85, 3.12) | 2.69% (2.57, 2.81) | * | * |
| Male/Female Sig |  |  |  |  |  |  |  |
| **Prevalence** |  |  |  |  |  |  |  |
| Both | 0.12% (0.03, 0.20) | 0.43% (0.35, 0.51) | * | 3.15% (3.05, 3.25) | 2.88% (2.78, 2.98) | * | * |
| Male | -0.04% (-0.12, 0.04) | 0.28% (0.20, 0.35) | * | 3.16% (3.07, 3.25) | 2.89% (2.80, 2.98) | * | * |
| Female | 0.24% (0.14, 0.34) | 0.54% (0.44, 0.64) | * | 3.11% (2.99, 3.22) | 2.84% (2.73, 2.95) | * | * |
| Male/Female Sig | * | * |  |  |  |  |  |

Mexican and USA border states exhibited divergent trends across metrics except deaths (p<0.05, Table 2). USA border states experienced rising DALYs (AAPC = 1.16%, 95% CI: 0.93, 1.39) and incidence (AAPC = 3.04%, 95% CI: 2.93, 3.16), while Mexican border states showed declining DALYs (AAPC = -0.62%) and incidence (AAPC = -0.15%). Despite these contrasts, mortality declined in both countries’ border states, with slightly larger decreases in the USA (AAPC = -1.28%, 95% CI: -1.83, -0.74) than Mexico (AAPC = -0.97%, 95% CI: -1.17, -0.76).

Significant sex differences in trends were observed across all geographies (Table 2). In Mexican border states, females had declining DALYs (AAPC = -1.05%, 95% CI: -1.21, - 0.90) and deaths (AAPC = -1.71%, 95% CI: -1.94, -1.48) while males showed minimal changes (all p<0.05). In Mexican non-border states, males had rising DALYs and deaths while females had declining trends. In USA border states, males had greater increases in DALYs (1.43% annually) and mortality than females (0.80% annually) (p<0.05, Table 2).

### 3.3 Joinpoint Regression Analysis (1990-2021)

Figure 1 presented joinpoint analysis of diabetes burden comparing border and non-border states in the USA and Mexico from 1990-2021. Within the USA, border states consistently outpaced non-border states in diabetes burden growth. The most rapid divergence occurred during 1995-2004, when border state incidence rose at 4.08% annually (95% CI: 3.86, 4.31) compared to 3.75% (95% CI: 3.55, 3.94) in non-border states. This gap persisted through 2021. DALYs showed similar patterns, with border states increasing faster both early (1990-2000: 3.37% vs 2.65%) and recently (2014-2021: 2.07% vs 1.57%). However, mortality trends were similar in both groups, declining significantly from 2001-2015 (border: APC=-3.91%; non-border: APC=-3.45%) before stabilizing. Mexico exhibited the reverse pattern. Border states stabilized or improved while non-border states deteriorated, particularly after 2009-2010. Non-border mortality reversed from declining (1990-2009: APC=-1.10%) to increasing (2009-2021: APC=1.49%, 95% CI: 0.97, 2.01). DALYs followed suit, shifting from improvement (1990-2010: APC=-0.64%) to deterioration (2010-2021: APC=1.27%, 95% CI: 0.84, 1.70). Border states showed no such reversal, maintaining stable or declining trends across metrics. However, Mexico border states had higher absolute burden across all metrics.

**Figure 1.**
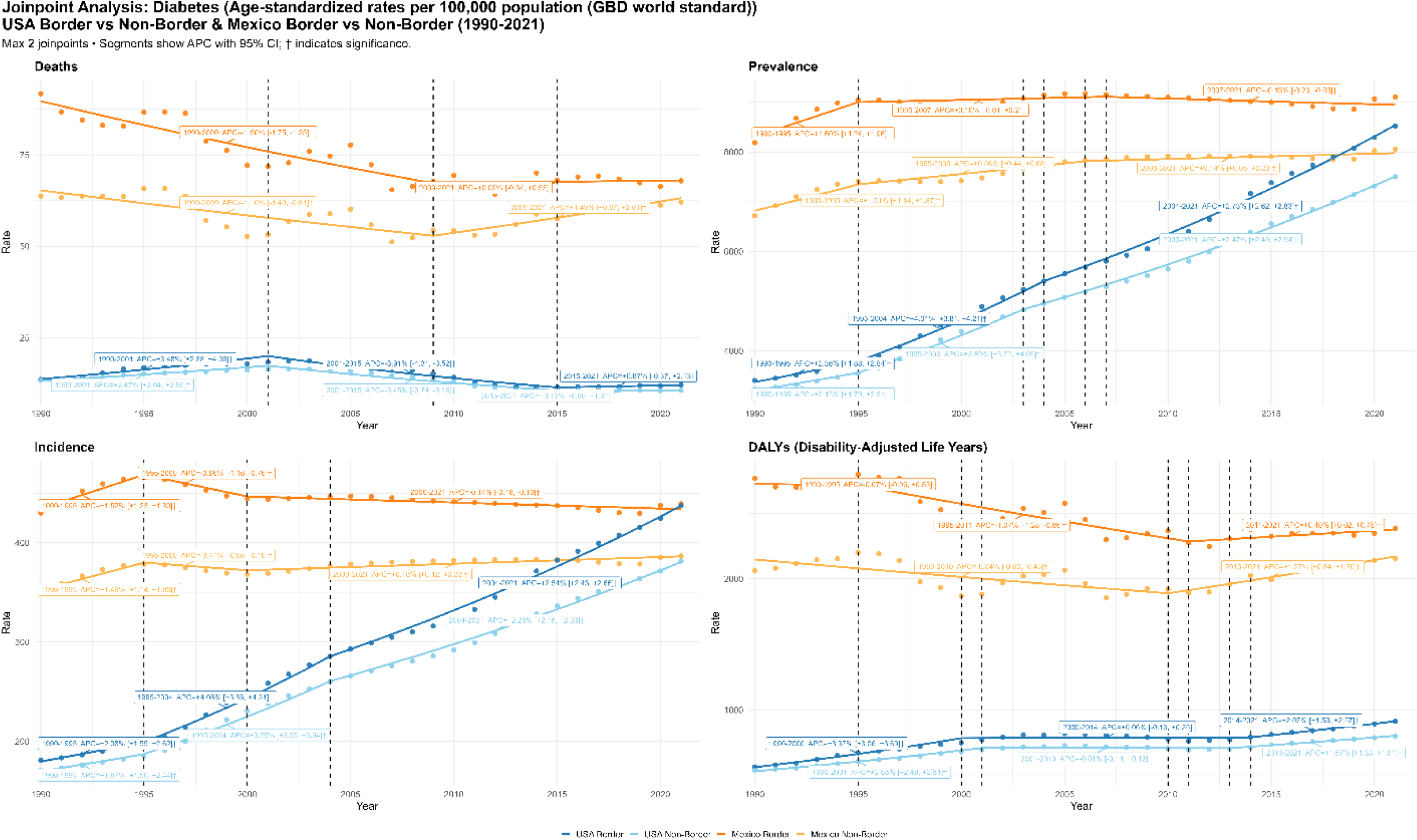
Joinpoint Regression Analysis of Diabetes Burden (1990-2021)

Figure 1 revealed a paradox between USA and Mexico border states. Mexico border states maintained substantially higher mortality (5-6 times higher) and DALY rates (3-5 times higher) throughout 1990-2021. However, prevalence and incidence converged, shifting from 2.4-fold differences in 1990 to near-parity by 2021.

### 3.4 Age-Period-Cohort Effect (1990-2021)

Figure 2 presented the age-period-cohort decomposition of diabetes rate ratios across four health metrics. The age-related patterns revealed consistent increases in diabetes burden across the lifespan, though with distinct peaks for different metrics. DALYs and prevalence both peaked around ages 60-70, reflecting the accumulated disability burden and disease prevalence in older adulthood, with Mexico Border states showing particularly high DALY ratios at these ages. Deaths increased exponentially with age. Incidence peaked sharply around age 60, particularly for USA border states, suggesting this is the critical age window for new diabetes onset, after which incidence declines.

**Figure 2.**
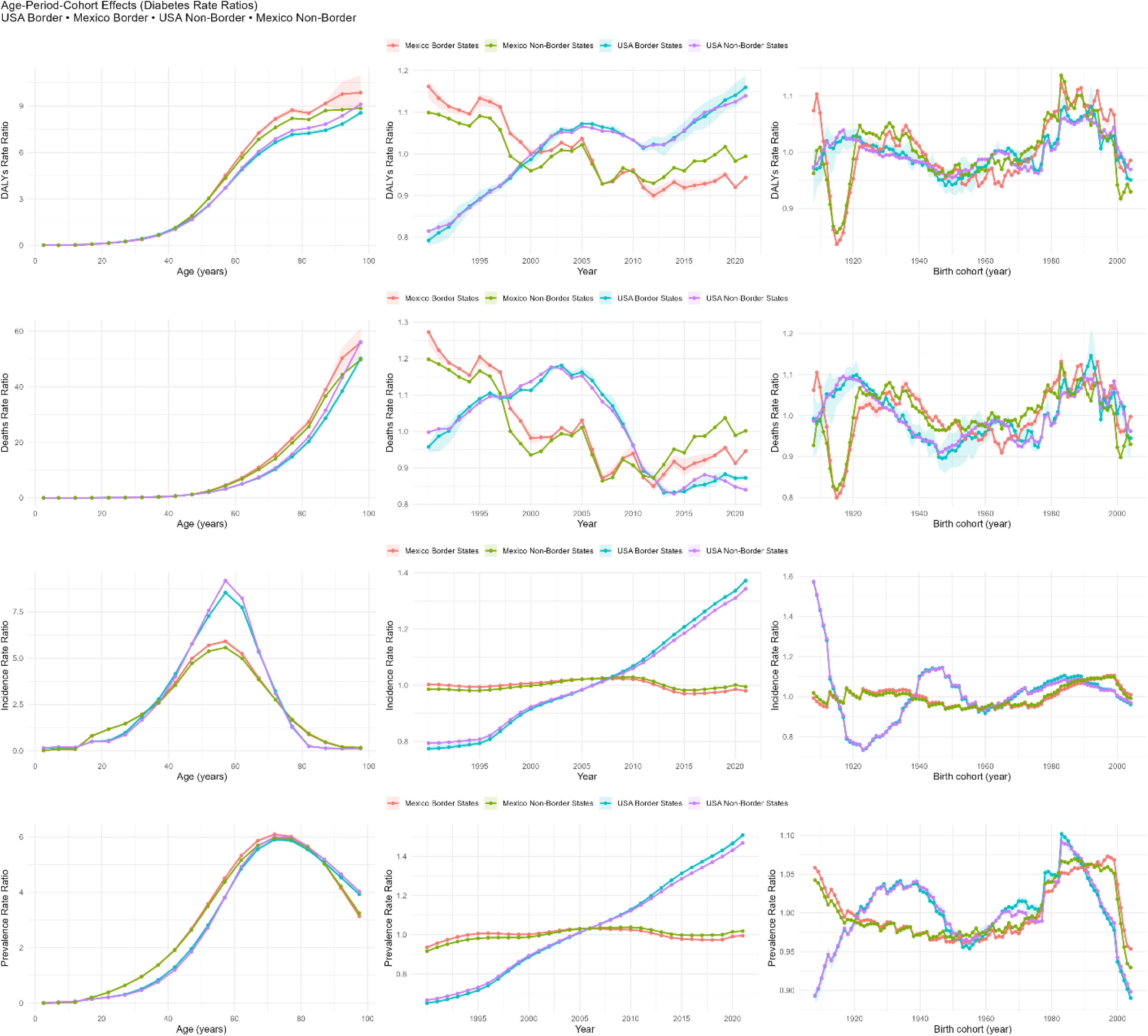
Age-Period-Cohort Effects for Diabetes Burden

Period trends revealed striking divergence between USA and Mexico border states despite shared geography. USA Border states showed increases across metrics from 1990 to 2021. DALYs rose from 0.8 to 1.15 (+44%), and incidence and prevalence climbed to 1.4-1.5. Conversely, Mexico Border states declined after 2005, with DALYs falling from 1.15 to 0.95 and deaths from 1.2 to 1.0, while incidence and prevalence remained flat around 1.0. Within the USA, border states consistently outpaced non-border states.

Cohort effects exhibited complex generational variations. DALYs showed a U-shaped pattern: dipping to 0.8 for the 1940 cohort, then rising to 1.1-1.2 for post-1980 births. Death ratios peaked at 1.05-1.15 for 1920-1940 cohorts, declined to 0.95-1.0 for mid-century births, then rose again to 1.05-1.10 for post-1980 cohorts. This indicated that recent generations faced 10-20% increased diabetes burden compared to mid-century cohorts.

### 3.5 Decomposition Analysis (1990-2021)

Figure 3 demonstrated divergent epidemiological transitions between USA and Mexico border states when comparing recent (2006-2021) versus early (1990-2005) periods. USA border states showed a dual pattern. For mortality indicators (DALYs, deaths, YLLs), substantial increases were driven entirely by population aging (orange bars) while underlying disease rates declined (negative blue bars). However, for morbidity indicators (incidence, prevalence, YLDs), large increases stemmed predominantly from rising disease rates rather than demographic shifts, indicating growing chronic disease burden independent of aging. Mexico border states displayed different patterns. Changes across all metrics were smaller in magnitude and driven primarily by aging effects, with disease rates remaining stable or declining. This resulted in minimal net change for deaths, YLLs, and YLDs. This contrasting decomposition revealed fundamentally different disease trajectories despite geographic proximity along the border. USA border states experienced rate-driven increases in diabetes burden despite mortality improvements, while Mexico border states maintained stable or declining rates across most metrics. Population growth is also a contributor to the increased burden of diabetes (Supplementary Figure 2).

**Figure 3.**
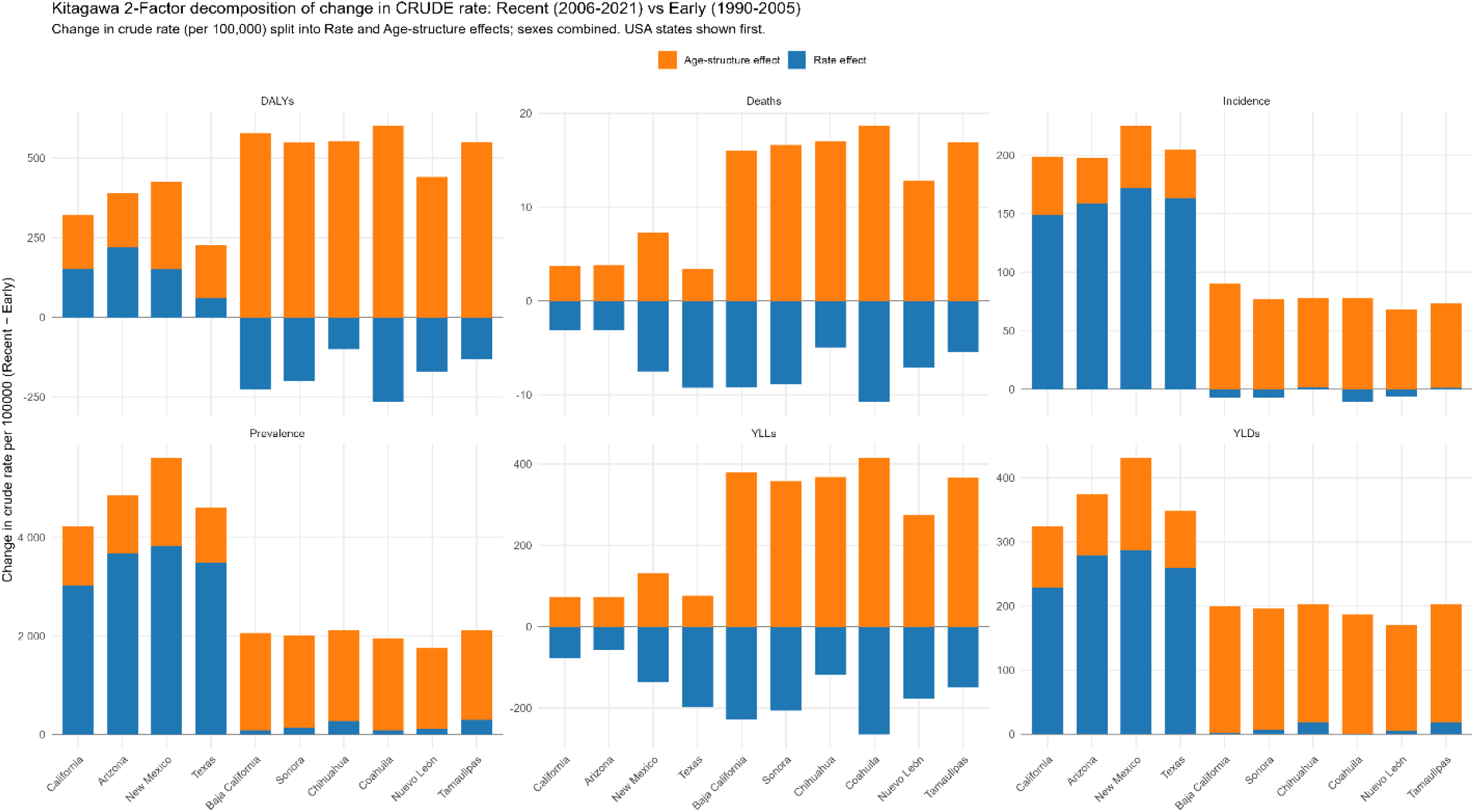
Diabetes Burden Decomposition Analysis for Border States

## 4. Discussion

This study provides the first comprehensive analysis of diabetes burden across USA-Mexico border states from 1990-2021, revealing a complex epidemiological landscape characterized by persistent cross-border health disparities despite converging disease occurrence. Diabetes Incidence (435 vs 425 per 100,000) and prevalence (9,010 vs 8,298 per 100,000) are near parity between Mexico and USA border states. However, mortality is nearly five-fold higher in Mexican border communities, and DALYs exhibit fundamentally different compositions. By integrating joinpoint regression, age-period-cohort models, and decomposition analyses, we uncover “epidemiologic cliffs” along the border.

In USA border states, incidence increases dramatically from 1990-2003 (AAPC +3.04%), reflecting rising obesity and improved detection, before plateauing. This high incidence, combined with substantially improved survival, drives continuous prevalence increases (AAPC +3.15%) even as mortality declines (AAPC -1.28%). The result is a growing population living with diabetes, “epidemiological success” where people survive with disease rather than dying from it. Mexican border states show the inverse pattern. Incidence declines modestly (AAPC -0.15%), likely reflecting persistent underdiagnosis rather than genuine reductions (Magliano et al., 2025). Prevalence increases minimally (AAPC +0.12%) because high mortality rates (AAPC -0.97%, plateauing after 2009) prevent accumulation of prevalent cases. This represents “epidemiological failure”, inability to transform an acute fatal condition into a manageable chronic disease.

USA border states show declining DALYs rates with YLDs share increasing from 42% to 68.6%, indicating successful shift from premature death to managed morbidity. Mexican border states show minimal DALYs decline with YLDs share reaching only 35.7%, meaning premature mortality (YLLs) still dominates 64% of burden. This incomplete epidemiological transition reflects healthcare systems that detect diabetes but cannot prevent its fatal complications (Gallardo-Rincón et al., 2023; OECD, 2016).

Global Burden of Disease 2021 analyses document that Mexican diabetes prevalence rose approximately 25% since 1990 with DALYs reaching 3.1 million in 2021 (6.6% of total), identifying high BMI as the dominant driver (Global Burden of Disease Collaborative Network, 2024).

Declining incidence in Mexican border states (-0.15% annually) versus rising then stabilizing incidence in USA border states (+3.04% until 2003) presents an epidemiological paradox. Given rising obesity in northern Mexico (Nájera & Ortega-Avila, 2024), true incidence declines seem implausible. Rather, persistent underdiagnosis in Mexico (Magliano et al., 2025) likely suppresses incidence estimates. If 30-40% of Mexican diabetes cases remain undiagnosed, reported incidence dramatically underestimates true occurrence. Advances in lowering undiagnosed cases influence both incidence and prevalence estimates (Fang et al., 2022), suggesting improved detection infrastructure would reveal substantially higher Mexican incidence. Sex-specific patterns support this interpretation. USA border states show male-predominant incidence (ratio 1.32) while Mexican border states show female-predominant patterns (ratio 0.86), likely reflecting differential case detection through women’s engagement with health services rather than true biological differences.

The mortality-to-prevalence ratio quantifies a crisis in diabetes management, 7.46 in Mexican border states versus 1.45 in USA border states. Mexican resident is approximately five times more likely to die from diabetes or its complications. This disparity persists across all age groups and both sexes, indicating systemic rather than population-specific failures. However, the true prevalence gap may be larger than our estimates suggest. Mexico experiences substantially higher rates of undiagnosed diabetes along with rising obesity trends in northern states, making it unlikely that Mexican diabetes prevalence is truly at parity with USA border states (Magliano et al., 2025; Nájera & Ortega-Avila, 2024). If 30-40% of Mexican diabetes cases remain undiagnosed (Magliano et al., 2025), the actual mortality-to-prevalence ratio becomes even more alarming. This means our analysis may underestimate the severity of Mexico’s diabetes management crisis.

The joinpoint analysis reveals when patterns diverged. USA prevalence accelerates sharply after 2003 (APC +2.73%), reflecting both rising incidence (APC +2.54%) and improving survival (i.e. declining deaths APC -3.91%). Mexican prevalence increases more modestly with no clear acceleration, suggesting detection improvements are offset by persistent high mortality. Mexico’s mortality improvements plateau after 2009, while USA mortality continues declining. This plateau has persisted for over a decade, suggesting Mexico has exhausted the benefits of prevention-focused interventions without building the management capacity to translate detection into survival (Gallardo-Rincón et al., 2023; OECD, 2016).

Sex-specific mortality patterns reveal additional complexity. Both countries show male-predominant mortality (ratios 1.71 in the USA, 1.12 in Mexico), but Mexico’s pattern combined with female-predominant diabetes occurrence (incidence ratio 0.86, prevalence ratio 0.82) suggests Mexican men with diabetes face catastrophic outcomes. This disconnect between occurrence and outcomes by sex in Mexico suggests differential healthcare access or utilization patterns, with men potentially more likely to be diagnosed but less likely to receive adequate management, including self-management resources. However, it is difficult to assume potential mechanisms for the increased mortality in men, without additional information about the main causes of death.

Age-specific analyses reveal that Mexico’s diabetes burden peaks earlier in life (ages 55-74) compared with the USA (ages 65-79), with mortality-to-prevalence ratios increasing more steeply with age. This premature burden in Mexico has profound implications for economic productivity as diabetes affects individuals during their most productive years. This impact on individuals is likely to result in reduced personal income and resources, which in turn amplifies challenges in diabetes management and increases health vulnerabilities for family members – particularly if the person with diabetes is the primary source of household income.

The decomposition analysis provides crucial insights into the drivers of changing diabetes burden. In USA border states, demographic factors (aging and population growth) drive increases in deaths and DALYs, while rate changes primarily drive prevalence increases. This pattern reflects successful mortality rate reductions being overwhelmed by demographic pressures and rising disease incidence. Conversely, Mexico shows modest rate improvements that partially offset demographic pressures, but these improvements remain insufficient to close the mortality gap with the USA. These findings suggest that different intervention strategies are needed on each side of the border. USA states must focus on primary prevention to reduce rising incidence rates, while Mexican states require urgent improvements in access to comprehensive diabetes management to reduce case fatality rates, even as they face similar demographic pressures (Bello-Chavolla et al., 2022).

Border-focused monitoring shows persistently higher burden within USA border states. Death rates increase by 9.7% from 2011-2018, with Texas border counties showing higher prevalence (15.5% versus 10.8%) and mortality (34.1 versus 21.4 per 100,000) than non-border counties (Alexander, 2020; Border Health Commission, 2024). The COVID-19 pandemic likely exacerbates vulnerabilities across all metrics through care disruptions, interrupted medication access, and detection disruptions (Bello-Chavolla et al., 2022; Gundlapalli, 2021).

### 4.1 Limitations

This analysis has several limitations that affect interpretation. First, relying on Global Burden of Disease 2021 estimates means inheriting modeling assumptions and uncertainty (Global Burden of Disease Collaborative Network, 2024). Second, Mexico’s higher undiagnosed rates mean true incidence and prevalence likely exceed estimates (Magliano et al., 2025). Third, comparing high-income and middle-income countries conflates healthcare infrastructure with true epidemiological differences, though disparity magnitude suggests real outcome differences. Fourth, state-level analyses mask within-state heterogeneity (*CDC WONDER*, 2024). Fifth, methodological choices, decomposition additivity assumptions, joinpoint selection, age-period-cohort identifiability constraints, influence the resulting estimates (Gupta, 1993; HORIUCHI et al., 2008; Irimata et al., 2022).

### 4.2 Implications

Our border-focused, multi-method analysis has clear implications for theory, practice, and policy. Theoretically, by separating population aging from true changes in diabetes rates, identifying when trends shift, and mapping age–period–cohort patterns, we extend life-course and place-based explanations and show that drivers differ across 1990–2005 and 2006–2021. Policy priorities include harmonized binational surveillance with shared metrics, sustained prevention and primary-care funding for border regions, population-level risk-reduction measures, and formal cross-border data sharing (Rosales et al., 2017). Overall, these results point to actionable, place- and cohort-informed strategies that target resources where they help most.

## 5. Conclusion

This first comprehensive binational analysis of diabetes burden across USA-Mexico border states (1990-2021) reveals a profound epidemiological paradox: converging disease occurrence alongside diverging outcomes. While incidence and prevalence have reached near-parity, Mexican border states maintain five-fold higher mortality and fundamentally different DALY compositions. USA border states demonstrate “epidemiological success”, transitioning diabetes from fatal to manageable (YLDs: 68.6% of burden), while Mexican border states exhibit “epidemiological failure”, unable to prevent fatal complications (YLLs: 64.3% of burden). The mortality-to-prevalence ratio of 7.46 versus 1.45 per 1,000 quantifies this crisis, likely underestimating severity given Mexico’s substantial undiagnosed cases. Despite geographic proximity and shared risk factors, disparities in diabetes outcomes persist across the border. Closing this gap requires binational commitment: primary prevention in USA states and urgent diabetes care management investment in Mexican states.

## Author Contributions

KW: design, analysis, interpretation, drafting, and editing. JH: review, and editing. CS: review, and editing. KK: review, and editing. GC: conception, interpretation, review, and editing.

## Data Availability Statement

The datasets generated and analyzed during the current study are available publicly as described in methods section.

---

**Supplementary Fig 1.**
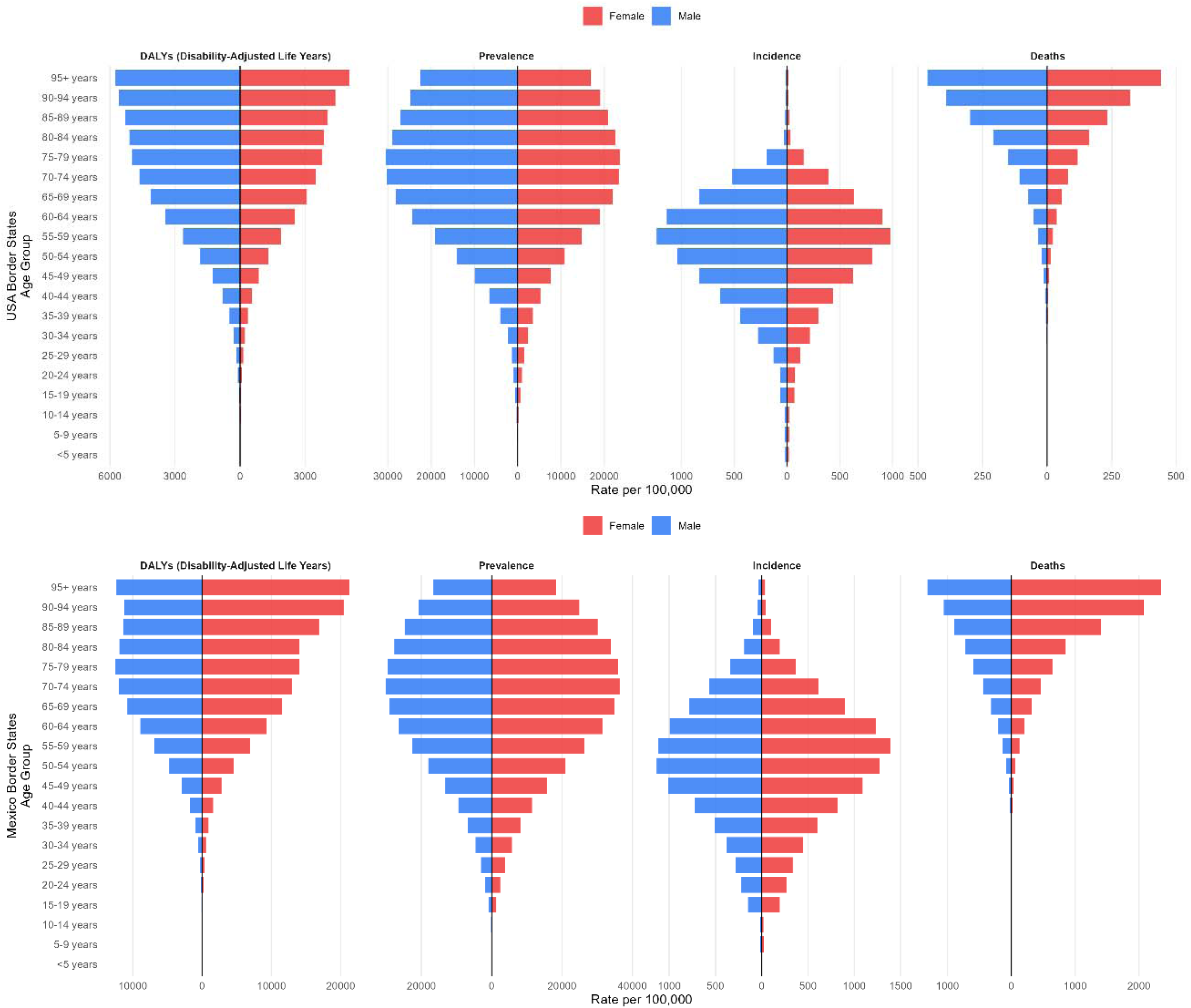
Age-Sex Distribution of Current Diabetes Burden in USA-Mexico Border States (2019-2021)

**Supplementary Fig 2.**
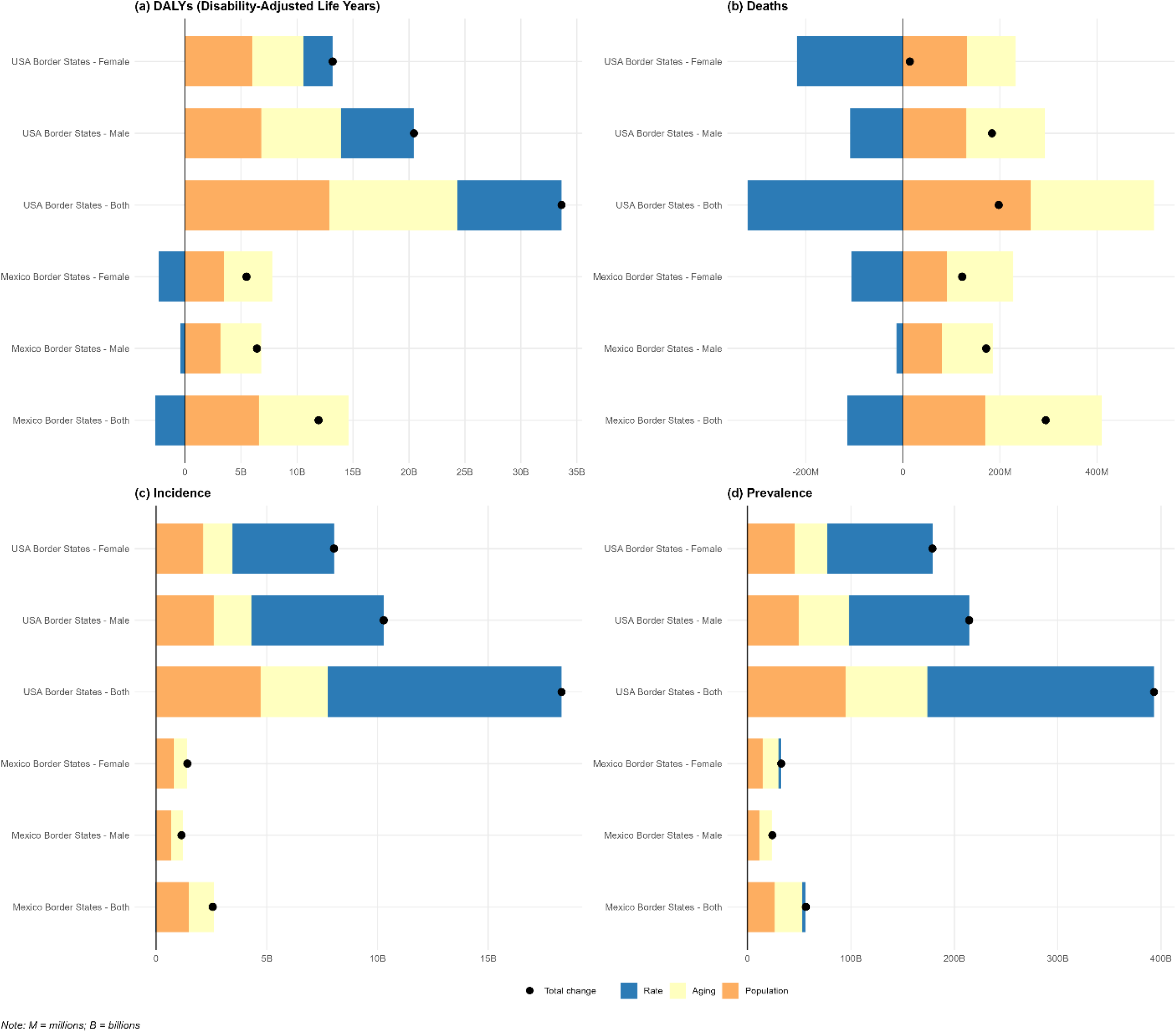
Decomposition by Sex for Border States by Das Gupta Method

