## Supplementary figures and images for "Closing the Gap: A Binational Analysis of Diabetes Mortality and Disability across the USA-Mexico Border Region"

### Supplementary Figure 1

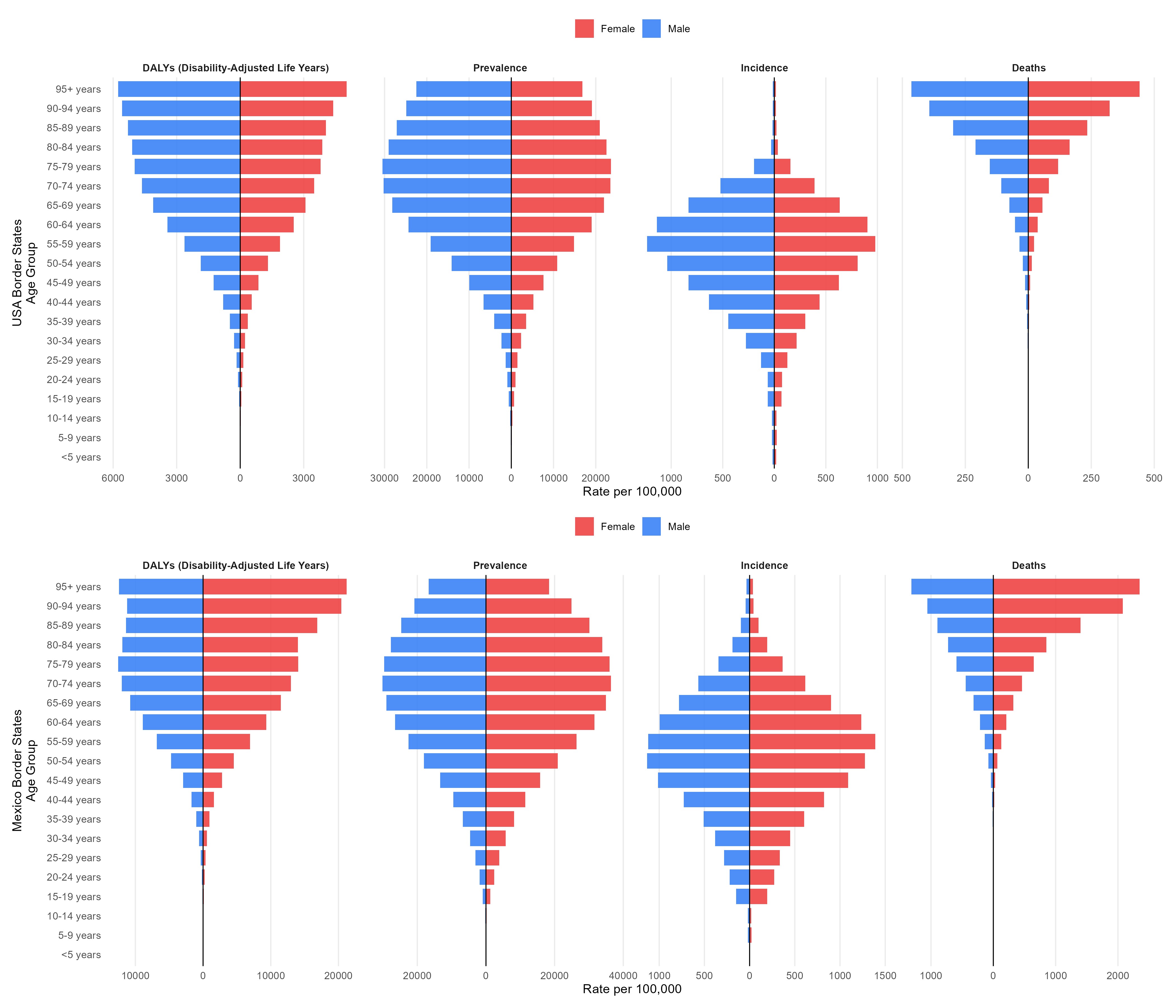

### Supplementary Figure 2

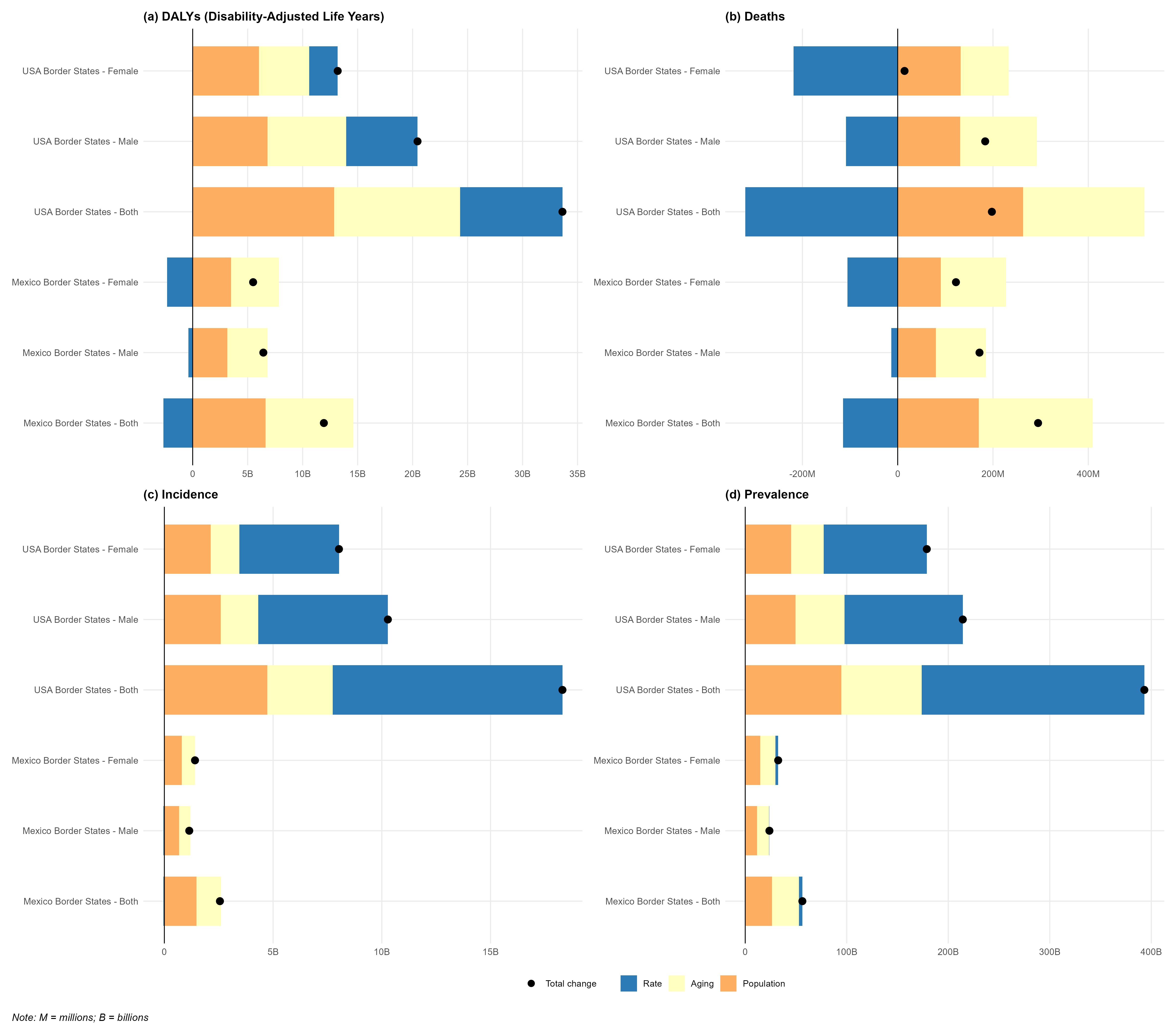
